# Economic Burden of Periodontal Disease in Europe and the United States of America – An updated forecast

**DOI:** 10.1101/2021.01.19.21250090

**Authors:** João Botelho, Luís Proença, Yago Leira, Leandro Chambrone, José João Mendes, Vanessa Machado

## Abstract

**Introduction:** Periodontal disease is a pandemic condition and its severe form affects approximately 10% of the global population. Here, we present a forecast for the economic burden of periodontal disease in 32 European countries and in the United States of America (USA).

**Material and methods:** In an aggregate population-based cost analysis, taken as reference the most recent available data, we estimated the cost of periodontal disease. Under this, global health, dental and periodontal expenditures were estimated. Additionally, indirect estimates accounted for Disability-Adjusted Life Years (DALY) valued at per capita Gross Domestic Product (GDP), to estimate productivity losses, including periodontal disease, edentulism due to periodontal disease and caries due to periodontal disease.

**Results:** In 2018 the aggregate cost in Europe was estimated at €17.00B and €2.35B more in the USA (€19.35B). Indirect costs due to periodontal disease amounted to €132.52B in European countries and €103.30B in the USA. The majority of the projected indirect costs were due to edentulism related to periodontal disease and periodontal disease. Indirect costs were the major portion of the estimated economic impact with an average of 0.66% of GDP in Europe and 0.50% in the USA. For the overall costs (direct and indirect), the value amounted to 0.75% of GDP in Europe and 0.60% in the USA.

**Conclusion:** Periodontal disease caused a €149.52B loss in Europe and €122.65B in the USA, in 2018. These results show that the economic burden of periodontal disease is increasing.

**CLINICAL RELEVANCE:** *Scientific rationale for the study:* Considering the pandemic pattern of periodontal diseases we present a forecast for the economic burden of periodontal disease in 32 European countries and in the United States of America (USA).

*Principal findings:* Periodontal disease caused a €149.52B loss in Europe and €122.65B in the USA, in 2018. For the overall costs (direct and indirect), the value amounted to 0.75% of GDP in Europe and 0.60% in the USA.

*Practical implications:* These results show that the economic burden of periodontal disease is increasing.

## 1. Introduction

Periodontal diseases are a major public health problem in Europe with significant socio-economic impacts (Hajishengallis, 2015; Kassebaum et al., 2014; Tonetti et al., 2017). Comprehensively, in 2010, severe periodontitis affected 10.8% people worldwide, while mild to moderate periodontitis affected the majority of the adult population and, therefore was considered the sixth-most prevalent health condition, worldwide (Kassebaum et al., 2014).

Health care expenditures have been increasing in the recent decades (Organisation for Economic Co-operation and Development [OECD] 2015). Given that the measurement of the economic burden associated with a disease is extremely useful to understand the maximum amount of resources that would gain if the disease were partially or totally eradicated (Rice, 1967). Building on such definition, highlighting the magnitude of the economic impact of periodontal disease in Europe and the United States of America (USA) allows to address oral public health measures by public health decision and policy makers.

Reporting the economic impact of oral diseases was a strategy previously used to describe data for 2010 (Listl et al., 2015) and 2015 (Righolt et al., 2018). In detail, both economic burden estimated direct costs associated with overall expenditures for dental health care and indirect costs connected to three most common oral conditions, namely untreated caries in permanent and deciduous teeth, severe periodontitis, and severe tooth loss (Listl et al., 2015; Righolt et al., 2018). Recently, the Global Burden of Disease (GBD) study predicted a direct cost of €44.28B ($54B) of periodontitis and €20.50B ($25B) in indirect costs in 2017 (James et al., 2018).

The expenditure associated with periodontal therapy is not only the treatment itself but also the indirect costs associated with the consequences of periodontal disease. In this context, 30% of root dental caries are associated with periodontal disease. Furthermore, the tooth loss as a negative consequence of severe periodontitis and its subsequent oral rehabilitation are also indirect costs. Nevertheless, it is important to point another non-clinical indirect cost, the productivity losses due to absenteeism from work. To the best for our knowledge, no study has comprehensively calculated the costs associated with periodontal disease.

Therefore, this study aimed to provide up-to-date estimates of economic burden (direct and indirect) of periodontal diseases across 32 European countries and compare it with the USA.

## 2. Methods

This economic burden study used a systematic approach to search and compute information regarding direct and indirect costs of periodontal diseases in each of the 32 European countries and the USA. The direct costs were defined as overall (public and private) expenditures for periodontal care. As indirect costs, we computed the productivity losses due to collateral costs associated with periodontal disease, in particular absence from work, impact on edentulism and impact on root caries.

Following a previously defined a priori protocol and with overall consensus among all authors, the most suitable methods were employed via detailed and best practice-based approaches. Considering that 2018 was the most recent year with a comprehensive set of data available, this year was defined as reference for estimation of the economic burden of periodontal disease. Also, this allowed us to line up with the GBD data for 2018. To this end, country-specific aggregate data from both national and international sources were gathered, following the approaches (Listl et al., 2015; Luengo-Fernandez et al., 2020).

### Estimation of direct costs

Country-specific aggregate health and dental expenditure data was sourced from: the WHO Global Health Expenditure Database (WHO, 2021). Annual Gross Domestic Product (GDP) was sourced from the Statistical Office of the European Communities (EUROSTAT) (*European Commission. Eurostat: Your Key to European Statistics*., n.d.), the International Labour Organization (ILOSTAT) (ILOSTAT, 2021) and the Organisation for Economic Cooperation and Development (OECD) Data (OECD, 2021).

The percentage of periodontal expenditures was not possible to obtain directly from each country. For this reason, we used information provided at the “National Services Scotland (NSS) Dental Statistics – NHS Treatment and Fees” of 2019, which has reported 23.3% of the overall treatment in the health system as being periodontal therapy (NSS, 2019). Thus, for each national dental expenditure, we considered 23.3% of the costs as associated with periodontal disease.

Taking into account that there was sufficient data for overall health and dental costs for all selected countries, it was not necessary to use data imputation techniques.

### Estimation of Indirect Costs

We followed the approach by Listl et al. (Listl et al., 2015), as per suggestion by the WHO’ s Commission on Macroeconomics and Health (WHO, 2001). In this approach, 1 Disability-Adjusted Life Year (DALY) was valued at one year of per capita GDP, to estimate productivity losses.

Country-specific data on health and dental expenditure was sourced from: the WHO Global Health Expenditure Database (WHO, 2021). To estimate country-specific data on mean daily earning we divided the mean annual earning by 200 (5 days of work per week x 50 weeks of work). Mean annual earning information was sourced from the EUROSTAT (*European Commission*, 2021), ILOSTAT, (ILOSTAT, 2021) and OECD data (OECD, 2021).

Country-specific data on the prevalence of periodontal disease cases were obtained from the most recent GBD study (GBD, 2021). These data were supplemented with individual patient-level data from national epidemiologic data.

To estimate edentulism due to periodontal disease, we arbitrarily attributed 50% of edentulism to periodontal disease according to previous estimates ranging from 35.8% (Ong, 1996, 1998) to 70% (EFP, 2021). For caries due to periodontal disease, we projected costs through a reported prevalence of 30% of caries patients having periodontal disease (Mattila et al., 2010), and then multiplying by a 82% prevalence of root caries in periodontitis patients (Reiker et al., 1999).

All data analyses were carried out with Microsoft Excel.

## 3. Results

### Direct costs

The overall estimation of dental health expenditure is set out in Table 1. The USA spent €3.5 trillion in health expenditures and Europe spent €2.0 trillion. In general, health expenditures represent an average of 8.40% of GDP in Europe, while in the US they represent 16.88%. The aggregate direct dental treatment costs in Europe were estimated at €88.96B, with 66.21% of the estimated expenditures (€48.29B) occurring in Germany, France, Italy, United Kingdom and Spain. Compared to the USA (€101.26B), European countries paid less €12.30B in the estimated health costs, and dental expenditures represent 0.4% of GDP while in the USA is 0.5%. Also, the dental expenditure cost paid out-of-pocket was €42.22B when compared to €40.50B in the USA.

**Table 1.** Costs of health, dental health and paid out-of-pocket (€ billion and % GDP) for 32 European countries and the United States of America in 2018.

| Country | Health expenditure (€B) | % GDP | Dental expenditure (€B) | % GDP | % Dental expenditure on Health expenditure | Dental expenditure cost paid out-of-pocket (€B) |
| --- | --- | --- | --- | --- | --- | --- |
| Austria | 46.99 | 10.33 | 2.71 | 0.6 | 5.77 | 1.41 |
| Belgium | 56.00 | 10.32 | 1.84 | 0.3 | 3.28 | 1.07 |
| Bulgaria | 4.86 | 7.35 | 0.20 | 0.3 | 4.01 | 0.19 |
| Croatia | 4.16 | 6.83 | 0.29 | 0.5 | 6.84 | 0.29 |
| Cyprus | 1.69 | 6.77 | 0.09 | 0.4 | 5.21 | 0.09 |
| Czech Republic | 18.73 | 7.65 | 0.95 | 0.4 | 5.08 | 0.50 |
| Denmark | 35.94 | 10.07 | 1.60 | 0.4 | 4.45 | 1.09 |
| Estonia | 2.05 | 6.69 | 0.20 | 0.6 | 9.61 | 0.15 |
| Finland | 24.94 | 9.04 | 1.19 | 0.4 | 4.75 | 0.81 |
| France | 313.86 | 11.26 | 13.61 | 0.5 | 4.34 | 2.99 |
| Germany | 453.06 | 11.43 | 30.52 | 0.8 | 6.74 | 7.63 |
| Greece | 16.83 | 7.72 | 0.81 | 0.4 | 4.82 | 0.81 |
| Hungary | 10.58 | 6.70 | 0.41 | 0.3 | 3.87 | 0.31 |
| Iceland | 2.20 | 8.47 | 0.15 | 0.6 | 6.73 | 0.11 |
| Ireland | 26.51 | 6.93 | 0.39 | 0.1 | 1.47 | 0.32 |
| Israel | 27.86 | 7.52 | 2.27 | 0.6 | 8.13 | 2.04 |
| Italy | 180.79 | 8.67 | 6.14 | 0.3 | 3.40 | 5.83 |
| Latvia | 2.13 | 6.19 | 0.13 | 0.4 | 6.05 | 0.12 |
| Lithuania | 3.51 | 6.57 | 0.32 | 0.6 | 9.18 | 0.26 |
| Luxembourg | 3.75 | 5.29 | 0.21 | 0.3 | 5.60 | 0.08 |
| Malta | 1.31 | 8.97 | 0.04 | 0.2 | 2.67 | 0.04 |
| Netherlands | 91.17 | 9.98 | 3.15 | 0.3 | 3.45 | 0.69 |
| Norway | 43.63 | 10.05 | 2.15 | 0.5 | 4.93 | 1.55 |
| Poland | 37.17 | 6.33 | 1.23 | 0.2 | 3.30 | 0.87 |
| Portugal | 22.80 | 9.41 | 0.77 | 0.3 | 3.40 | 0.76 |
| Romania | 13.42 | 5.56 | 0.45 | 0.2 | 3.37 | 0.41 |
| Slovakia | 7.08 | 6.69 | 0.33 | 0.3 | 4.72 | 0.12 |
| Slovenia | 4.48 | 8.30 | 0.22 | 0.4 | 4.93 | 0.04 |
| Spain | 127.67 | 8.98 | 4.34 | 0.3 | 3.40 | 4.25 |
| Sweden | 60.54 | 10.90 | 3.37 | 0.6 | 5.56 | 2.05 |
| Switzerland | 83.74 | 11.88 | 4.62 | 0.7 | 5.52 | 3.60 |
| United Kingdom | 291.58 | 9.98 | 4.29 | 0.1 | 1.47 | 1.74 |
| Europe (Total) | 2.021.03 | 8.40 | 88.96 | 0.4 | 2.91 | 42.22 |
| United States of America | 3.475.02 | 16.88 | 101.26 | 0.5 | 5.77 | 40.50 |

Concerning the direct costs of periodontal disease, the aggregate cost in Europe in 2018 was estimated at €17.00B and €2.35B more in the USA (€19.35B). Largely, Germany, France, Italy, United Kingdom and Spain represented the majority of the projected expenditures.

### Indirect Costs

The estimates of indirect costs, in other words productivity losses, are presented in Table 2. Indirect costs due to periodontal disease amounted to €132.52B in European countries and €103.30B in the USA. The majority of the projected indirect costs were €72.71B (54.9%) because of edentulism due to periodontal disease, €55.68B (42.0%) to periodontal disease and €4.14B (3.1%) to untreated caries in permanent teeth due to periodontal disease (root caries). The highest productivity losses were found for Germany (€29.55B), France (€19.28B), Italy (€14.65B), United Kingdom (€14.26B), Netherlands (€7.60B) and Spain (€6.00B). In the USA, the majority of the projected indirect costs were also due to edentulism, related to periodontal disease (€50.42B, 48.8%) and periodontal disease (€41.13B, 39.8%).

**Table 2.**
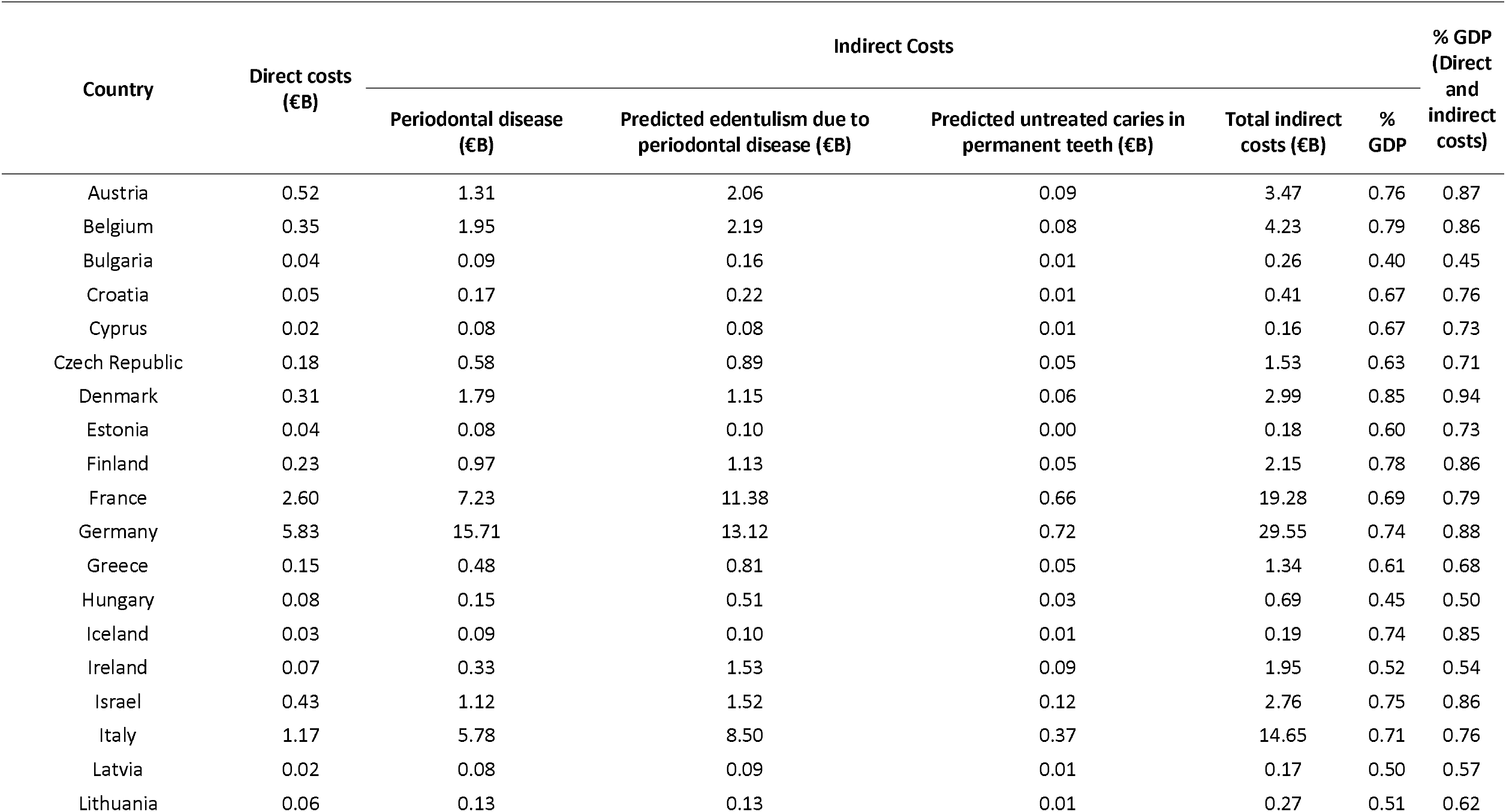
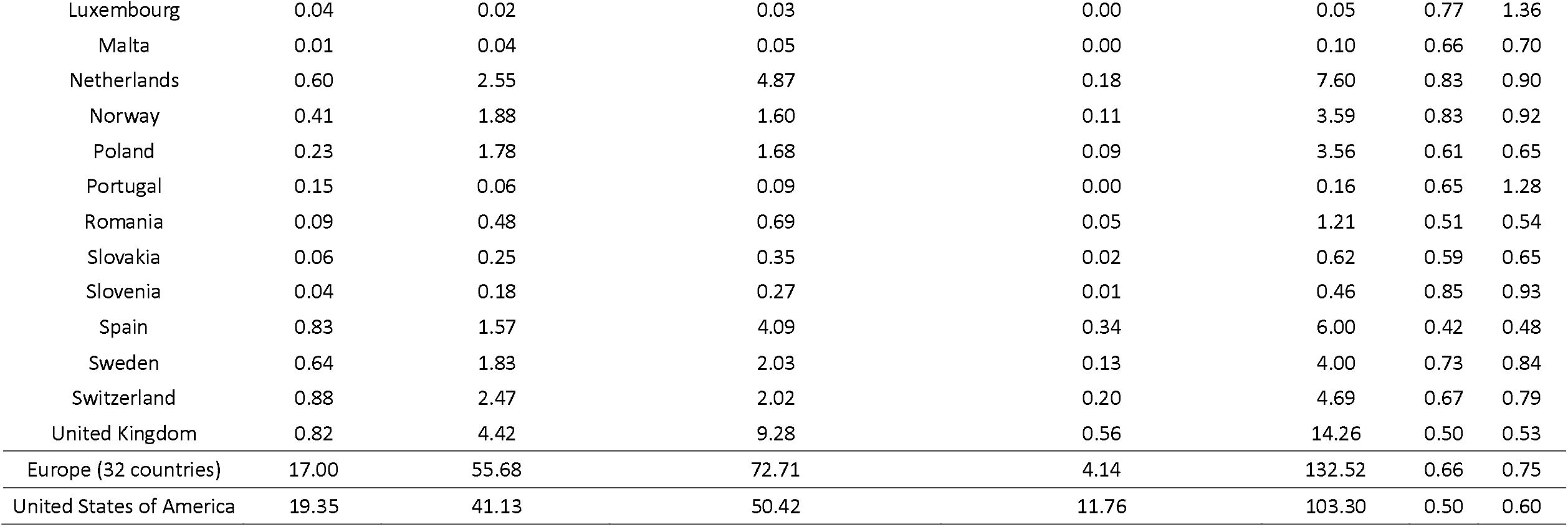
Estimated direct and indirect cost by periodontal disease (€ billion and % GDP) for 32 European countries and the United States of America in 2018.

As with the impact on GDP, indirect costs were indeed the majority of the estimated economic impact with an average of 0.66% of GDP in Europe and 0.50% in the USA. In the overall costs (direct and indirect), the value amounted to 0.75% of GDP in Europe and 0.60% in the USA. Combining direct and indirect projected costs, the total impact of periodontal disease was €149.52B in Europe and €122.65B in the USA.

## 4. Discussion

We estimated the total cost of periodontal disease, in 2018, across 32 European countries to be approximately €149.52B. From these forecasts, the majority were incurred by indirect costs due to periodontal disease, which amounted to 0.66% of the European GDP. Among the indirect costs, periodontal disease and edentulism due to periodontal disease were the most impactful determinants.

In the USA, the expenditure of periodontal disease was projected at €122.65B in 2018, less €26.87B than Europe. Furthermore, this prediction calculated the impact of indirect costs at 0.50% of the USA GDP, and 0.60% for the total costs. Similar to Europe, periodontal disease and edentulism due to periodontal disease were the most impactful indirect costs estimated.

Mindful on the burden of health expenditure in the total GDP, the USA invested closely twofold the percentage of health expenditure than European countries (16.88% vs. 8.40%). Seemingly, the indirect cost of periodontal disease was lower in the USA. Although we cannot deduce an association, a possible preventive attitude towards periodontal diseases may be a reason for this result. To this end, the USA Office of Disease Prevention and Health Promotion, via Centre for Diseases Control evidence, set the ambition to reduce the prevalence of periodontal disease by 6.7% (*U. S. Department of Health and Human Services Healthy People 2020*., 2010). Ever since, the detection and preventive action towards periodontal disease has been extremely forceful (Eke et al., 2016, 2018, 2020), and these may explain such lower impact on indirect costs, compared to the European scenario. Furthermore, such vigorous action has been carried out by the European Federation of Periodontology (Caton et al., 2018; Sanz et al., 2020), and one could say reaching this level of impact may require a broad collaboration with European regulatory organizations (i.e. the European Centre for Disease Prevention and Control).

Regarding the precision of our results, the quality and availability of data are decisive to attain comparable periodontal disease-related data across countries (Burns et al., 2016; Leal et al., 2016). Considering that thirty-three countries were analyzed (32 European countries and the USA), we required a number of sources and have experienced insufficiencies in available information to compute costs, and for this reason assumptions had to be made. Such approach is common and has been previously reported (Burns et al., 2016; Leal et al., 2016; Luengo-Fernandez et al., 2013, 2016, 2020). Yet, both health and dental expenditures were mostly available in European official databases which increase the reliability of these results. As the 2019 data is still scarce and estimates by regional comparison with socioeconomically similar countries would have to be made, we decided to focus on the 2018 data. Another important fact is the currency conversion, since we converted dollars to euros as we planned this report towards the European setting, and this may have contributed with imprecisions to the final estimates. Nevertheless, these projections are novel and demonstrate a growing economic burden of periodontal disease in Europe compared to 2010 estimates (Listl et al., 2015).

Importantly, the anticipated direct costs due to periodontal disease are inexact as national statistics on the incidence of periodontal treatments are seldom. Thus, we decided to use data from the “National Services Scotland Dental Statistics – NHS Treatment and Fees” of 2019, that reported 23.3% of treatment being periodontal therapy (NSS, 2019). Indeed, this result presents some limitation, however future national information is crucial to estimate precisely the direct costs involved with periodontal disease. As for the indirect cost, the approach used has been validated in the past (WHO, 2001).

This study presents some other limitations worth mentioning. There can be some level of underestimation because some categories directly associated with periodontal disease were not recorded as health statistics (such as, periodontal surgical therapy, implant therapies after tooth extraction due to periodontitis, peri-implantitis therapies, orthodontic treatments due to periodontal disease or restorative dentistry to mitigate soft tissue esthetic defects). These categories of expenditure were not included due to scarcity of data and for this reason we highlight the need for further collection in the future. Also, because of this hypothetical lack of information, and as far as we may estimate, these overall costs may be significantly superior.

Lastly, these results not only present disturbing evidences about the undeniable impact that periodontal disease has on the economy, but they also show that it has been increasing since 2010. Aware of the time we live in, with the COVID-19 pandemic, worrisome economic decline and consequently less financial capacity, health in general, and consequently oral health (including periodontal) will deteriorate. For this reason, we can conjecture that periodontal disease may grow even more with presumable higher economic impact. More than ever, it is fundamental to reinforce periodontal public health measures and articulate with national and European entities.

## 5. Conclusion

Our report offers a view of the economic costs posed by periodontal disease to 32 European countries and in the USA in 2018. Periodontal disease caused a €149.52B loss in Europe and €122.65B in the USA that year. These results show that the economic burden of periodontal disease is increasing.

## Data Availability

Data will be provided upon reasonable request

## Informed consent

Not applicable.

## Ethical approval

Not applicable.

## Acknowledgements

Yago Leira has a research contract with Fundación Instituto de Investigación Sanitaria de Santiago de Compostela (fIDIS) and holds a Senior Clinical Research Fellowship supported by the UCL Biomedical Research Centre who receives funding from the NIHR (NIHR-INF-0387).

